# Adverse childhood experiences, social support, and cardiovascular-kidney-metabolic syndrome: evidence from two national cohorts

**DOI:** 10.1101/2025.09.11.25335611

**Authors:** Xiu Qin, Haofeng Zhang, Fudong He, Lei Sun, Hua Xiao, Yufan Nan, Haidong Zhu, Yanbin Dong, Haiyan Chen, Guang Hao

**Affiliations:** School of Public Health, Guangdong Pharmaceutical University, Guangzhou, China; School of Health, Guangdong Pharmaceutical University, Guangzhou, Guangzhou, China; Georgia Prevention Institute, Department of Medicine, Medical College of Georgia, Augusta University, Augusta, Georgia, USA; Guangzhou Center for Disease Control and Prevention (Guangzhou Health Supervision Institute), Guangzhou, China

**Keywords:** social support, adverse childhood experiences, cardiovascular-kidney-metabolic syndrome

## Abstract

**Background:** The association between adverse childhood experiences (ACEs) and cardiovascular-kidney-metabolic (CKM) syndrome, as well as the role of social support in this association, has not been elaborated.

**Methods:** Participants with complete information from the Health and Retirement Study (HRS) and the China Health and Retirement Longitudinal Study (CHARLS) were included. In the main analysis, CKM stages 3 - 4 were classified as advanced CKM, and logistic regression was used to analyze the relationship between ACEs and advanced CKM. Pooled effects were determined through random-effects meta-analysis, with Cochran’s Q test value and I^2^ statistic reported to assess heterogeneity. Mediation analysis was performed to investigate the mediating role of social support in this association.

**Results:** The study included 6100 and 9722 eligible participants from the HRS and CHARLS, respectively. After adjusting for covariates, compared to the participants without ACEs, those who experienced more ACEs had a higher risk of advanced CKM (in participants who experienced 1 ACEs: pooled OR = 1.12, 95%CI:0.95 - 1.32, *P* = 0.168; 2 ACEs: pooled OR = 1.25, 95%CI:1.11 - 1.41, *P* < 0.001; ≥ 3 ACEs: pooled OR = 1.31, 95%CI:1.18 - 1.45, *P* < 0.001). Additionally, social support mediated 2.1% and 11.8% the association between ACEs and CKM syndrome in the CHARLS and HRS cohorts, respectively.

**Conclusion:** We for the first time reported that ACEs were significantly associated with CKM syndrome in the general population. Furthermore, our results suggest that social support plays a partial mediating role in this relationship.

## Introduction

Adverse childhood experiences (ACEs), refer to a range of negative life events that occur before the age of 18, have become a significant public health concern worldwide.^1-3^ The adverse health outcomes of ACEs are multifaceted and affect individuals throughout their lifespan. Current evidence suggests that ACEs increase the risk of cognitive impairment,^4^ dementia,^5^ cardiovascular diseases (CVD),^6^ and cancer.^7^ The American Heart Association (AHA) has recently proposed the notion of CKM syndrome to address the complex interactions between CVD, chronic kidney disease, and metabolic disorders in 2023.^8^ It encompasses a spectrum of comorbidities, including obesity, diabetes, hypertension, dyslipidemia, and chronic kidney disease, which often coexist and contribute to adverse health outcomes.^9^ A recent study reported that the prevalence of CKM in the US was 90% (stage 1 or higher) and 15% met criteria for advanced stages(stage 3 or 4) in 2020.^10^ In China, nearly 80% of Chinese adults met criteria for CKM syndrome, and 23.6% were advanced CKM.^11^ Although some studies reported the associations between ACEs and CKM components, the relationship between ACEs and CKM has not been studied.

Social support has been defined as any “support accessible to an individual through social ties to other individuals, groups, and the larger community”.^12^ The protective role of social support in mitigating the adverse effects of stress has been well-documented. The model proposed by Cohen and Wills highlights how social support can buffer stress through both structural aspects, such as the presence of relationships, and functional aspects, such as perceived support in addressing stress-related needs.^13^ Research has found that complementing ACEs prevention efforts with social support interventions could be an effective strategy to reduce premature deaths among US young adults, and confirmed partial mediating effects of social support on the relation between ACEs and anxiety and ACEs and depression. ^14,15^ Additionally, higher levels of social support have a protective effect in individuals with or without CVD.^16,17^

We hypothesize that ACEs were associated with a higher risk of CKM, and social support plays a role in this association. Here, we investigated the relationship between ACEs and CKM using two national longitudinal cohorts, as well as the mediating role of social support in the association. The results can shed light on the earlier interventions and tailored strategies for reducing the risk of CKM.

## 2 Method

### 2.1 Study population

The data used in this study came from two nationally longitudinal cohorts: the Health and Retirement Study (HRS, https://hrs.isr.umich.edu/) and the China Health and Retirement Longitudinal Study (CHARLS, https://charls.pku.edu.cn/en/), all of which followed the ethical guidelines outlined in the Declaration of Helsinki. The HRS, launched in 1992, is a comprehensive study in the US that tracks various aspects of ageing every two years, including health, work, retirement, social connections, and economic status.^18^ It adheres to the University of Michigan’s Institutional Review Board guidelines and has informed consent from participants. The CHARLS is a nationally representative micro-database in the households of the elderly and individual information on elderly respondents and their spouses, including health, economy, and retirement etc. The detailed study design and sampling methods have been reported previously. The CHARLS, initiated in June 2011, surveys a nationally representative sample of Chinese aged 45 and older, with follow-ups every two to three years, approved by Peking University’s Ethical Review Committee, and requires informed consent from all participants before participation.^19^

In this study, we used data from the CHARLS conducted from June to December 2014, and CKM syndrome individuals who were interviewed in Wave 3 in 2015. The data from HRS were used in Wave 2015 - Wave 2019. The exclusion criteria of the two cohorts were as follows: 1) with missing information on life history questionnaires, covariates 2) with missing information on CKM individuals.) Finally, there were 6100 and 9722 eligible respondents from the HRS and the CHARLS available for analysis, respectively. (**Supplementary Figure 1**)

### 2.2 Adverse childhood experiences

In the HRS, ACEs were defined as eight items: 1) having both parents with less than a high school education, or one parent with less than a high school education if only one parent was reported, 2) experiencing physical abuse from a parent, 3) exposure to parental alcohol and drug use before age 18, 4) separation from either parent during childhood, 5) death of a parent, 6) residing in a children’s home or orphanage, 7) childhood poverty, and 8) parental separation or divorce before age 16.^20^ Specifically, due to a large-scale missing on parental education, it was excluded from the main analysis. In the CHARLS, ACEs included 12 items: 1) physical abuse, 2) emotional neglect, 3) substance abuse within the household, 4) mental illness in the household, 5) domestic violence, 6) having an incarcerated household member, 7) parental separation or divorce, 8) living in an unsafe neighborhood, 9) being bullied, 10) death of a parent, 11) death of a sibling, and 12) parental disability.^21^

All items related to ACEs in the two cohorts were binary, and each respondent received 1 point for each condition reported, with a total score ranging from 0 to 7 in the HRS and from 0 to 12 in the CHARLS. Additionally, participants were categorized into 4 groups according to their ACE scores: 0, 1, 2, and ≥ 3. The details of each ACE item of the two cohorts were shown in **Supplementary Table S1.**

### 2.3 Cardiovascular-Kidney-Metabolic Syndrome

According to the guideline, 5 stages of CKM syndrome were assessed: stage 0 (no CKM risk factors), stage 1 (excess and/or dysfunctional adiposity), stage 2 (metabolic risk factors and CKD), stage 3 (subclinical CVD), and stage 4 (clinical CVD). CKM Stage 0 included participants with normal BMI (< 23 kg/m^2^) and normal WC (< 80 cm for women or < 90 cm for men). Participants meeting the criteria of the elevated stages were diagnosed as higher stages. CKM Stage 1 identified individuals with elevated BMI (≥ 23 kg/m^2^), elevated waist circumference (≥ 80 cm for women or ≥ 90 cm for men), or prediabetes. Prediabetes was diagnosed through fasting blood glucose (100 to < 126 mg/dL) or glycated hemoglobin (5.7% to < 6.5%). CKM Stage 2 identified participants with metabolic risk factors, moderate-to-high-risk CKD (estimated glomerular filtration rate [eGFR] 45 to < 60 mL/min/1.73 m^2^), or both. eGFR was calculated using the race-free CKD-EPI 2021 creatinine equation. Metabolic risk factors included elevated fasting serum triglycerides (≥ 135 mg/dL), hypertension, diabetes, or metabolic syndrome. The metabolic syndrome was identified by the presence of 3 or more of the following conditions: elevated waist circumstance, low high-density lipoprotein cholesterol levels (< 40 mg/dL for men or < 50 mg/dL for women), elevated triglyceride levels (≥ 150 mg/dL), high BP (systolic BP *≥* 130 mm Hg or diastolic BP *≥* 80 mm Hg), or prediabetes. CKM Stage 3 was identified based on the presence of very-high-risk CKD stages (eGFR 15 to < 45 mL/min/1.73 m^2^), or a self-reported CKD, or a high-predicted 10-year CVD risk (≥ 20%). The 10-year cardiovascular risk was estimated with the American Heart Association Predicting Risk of CVD Events (PREVENT) equations. CKM Stage 4 was identified based on self-reported established CVD (coronary heart disease, angina, heart attack, heart failure, and stroke) among individuals with excess / dysfunctional adiposity, other metabolic risk factors, or CKD.^22^ Stages 3 and 4 are collectively classified as advanced CKM syndrome, encompassing individuals diagnosed with or at high risk of developing cardiovascular disease.^10^

### 2.4 Social support

In the HRS, three items assessing social support include: ‘‘How much do they really understand the way you feel about things?’’ ‘‘How much can you rely on them if you have a serious problem?’’ and ‘‘How much can you open up to them if you need to talk about your worries?’’ Items were asked in four loops in reference to participants’ spouse / partner, children, family members, and friends. The response options ranged from 1 (a lot), 2 (some), 3 (a little), to 4 (not at all). Items were re-coded so that a higher value indicates a higher level of social support. Social support from each of the four sources was calculated separately by the average of the above three items measuring support from the corresponding source.^23^ Young adulthood social support, encompassing economic, non-economic, and emotional support, was also assessed in the CHARLS dataset through the following questions: “When you were a young adult, was there anyone who provided you with financial support for your work?”, “When you were a young adult, was there anyone who provided you with positive nonfinancial support for work?”, and “when you were a young adult, was there anyone who provided you with positive support or mentoring for your interpersonal relationship?”. Responses were dichotomized, and a total score ranging from 0 to 3 was calculated, with higher scores indicating greater social support.^24^ The details of each social support item of the two cohorts were shown in **Supplementary Table S2.**

### 2.5 Covariates

Covariates included age, sex (male or female), marital status (married or unmarried), residential area (rural or urban), Current smokers were those who “smoke cigarettes now”, and current drinkers referred to participants who consumed alcohol in the past three months. For the HRS, high-level education included high school diplomas and above (≥ 12 years). In the CHARLS, high-level education encompassed junior high school diplomas and above (≥ 9 years).

### 2.6 Statistical analysis

Continuous variables were reported as means ± standard deviation (x̄ ± s), while categorical variables were given as percentages (n [%]). A logistic regression model was employed to investigate the longitudinal relationship between ACEs and advanced CKM syndrome. It adjusted for age, sex, marital status, education level, smoking, drinking, race (white / black, available in the HRS), and residential area (rural / city, available only in the CHARLS). Pooled effects were determined through random-effects meta-analysis, and heterogeneity was assessed with Cochran’s Q test and I^2^ statistic. Stratified analyses were conducted based on sex (male vs. female), education level (low vs. high), smoker (yes or no), drinker (yes or no), race (White / Black, available in the HRS), residential area (rural / city, available only in the CHARLS), and median age (for the CHARLS cohort: < 62 years vs. ≥ 62 years; for the HRS cohort: < 68 years vs. ≥ 68 years). To evaluate potential effect modifications, multiplicative interaction terms between these factors and ACEs were included in the logistic regression models. In R software (Version 4.4.1), the "mediation" package was used for mediation analysis. Indirect, direct, and total effects were estimated using bootstrap resampling (1000 iterations), and 95% confidence intervals were calculated. In a sensitivity analysis, the associations of individual ACEs with advanced CKM were assessed. A linear regression model was employed to investigate the relationship between ACEs and CKM syndrome in another sensitivity analysis. All analyses were performed with Stata 18.0 (Stata Corp., TX, US) and R (Version 4.4.1). A two-sided *P* < 0.05 was considered statistically significant.

## 3 Results

### 3.1 General characteristics

The analysis included 6100 participants from the HRS and 9722 from the CHARLS. The average age was 68.50 ± 10.70 years for the HRS participants and 62.27 ± 9.09 years for the CHARLS participants. Female participants accounted for 51.03% and 53.48%, respectively. Most individuals experienced at least one ACE, with 20.93% in the CHARLS and 20.62% in the HRS experiencing ≥ 3 ACEs. There were 35.09% participants who had advanced CKM (stage 3 - 4) in the CHARLS cohort, and 38.29% had advanced CKM in the HRS. (**Table 1**) The distribution of ACEs in two cohorts was presented in **Supplementary Figure 2**.

**Table 1.** Characteristics of the two cohorts.

| Characteristics | CHARLS | HRS |
| --- | --- | --- |
| No. | 9722 | 6100 |
| Age, year | 62.27±9.09 | 68.50±10.70 |
| Female, % | 5199(53.48) | 3113(51.03) |
| Married, % | 8331(85.69) | 5741(94.11) |
| High-level education, % | 5211(53.60) | 4758(78.00) |
| Current drinker, % | 3332(34.27) | 3840(62.95) |
| Current smoker, % | 4329(44.53) | 1278(20.95) |
| Urban/White % | 1801(18.52) | 4481(73.46) |
| Number of ACEs, % |  |  |
| 0 | 2561(26.34) | 2147(35.20) |
| 1 | 2875(29.57) | 1639(26.87) |
| 2 | 2251 (23.15) | 1056(17.31) |
| ≥3 | 2035(20.93) | 1258(20.62) |
| Number of CKM syndrome, % |  |  |
| 0 | 624(6.42) | 127(2.08) |
| 1 | 1772(18.23) | 1416(23.21) |
| 2 | 3915(40.27) | 2221(36.41) |
| 3 | 1289(13.26) | 693(11.36) |
| 4 | 2122(21.83) | 1643(26.93) |

Abbreviations: ACEs: adverse childhood experiences;
CKM syndrome: cardiovascular-kidney-metabolic syndrome; CHARLS: China Health and Retirement
Longitudinal Study; HRS: Health and Retirement Study; SD: standard deviation. Continuous variables were reported as means ± standard deviation, while categorical variables were given as percentages (n [%]).

### 3.2 Association between ACEs and CKM syndrome

The proportions of advanced CKM were 32.88%, 33.88%, 36.47% and 38.03% in the participants who experienced 0, 1, 2, and ≥ 3 ACEs in the CHARLS cohort, respectively. The corresponding figures in the HRS cohort were 35.72%, 42.16%, 39.96%, and 36.25%. **(Figure 1)** Adjusted for other risk factors, in the CHARLS cohort, compared with those who did not have ACEs, the risk of advanced CKM in participants with ACEs of 2 and ≥ 3 increased 19% (OR = 1.19, 95%CI:1.05 - 1.35, *P* = 0.007) and 34% (OR = 1.34, 95%CI: 1.18 - 1.52, *P* <0.001), respectively. Similarly, in the HRS cohort, compared with those who did not have ACEs, the risk of advanced CKM in participants with ACEs of 1, 2 and ≥ 3 increased 23% (OR = 1.23, 95%CI: 1.06 - 1.44, *P* = 0.007) a 35% (OR = 1.35, 95%CI: 1.13 -1.61, *P* = 0.001) and 34% (OR = 1.34, 95%CI: 1.06 - 1.49, *P* =0.008), respectively. Further, compared to the participants without ACEs, those who experienced more ACEs had a higher risk of advanced CKM (in participants who experienced 1 ACEs: pooled OR = 1.12, 95%CI:0.95 - 1.32, *P* = 0.168; 2 ACEs: pooled OR = 1.25, 95%CI:1.11 - 1.41, *P* < 0.001; ≥ 3 ACEs: pooled OR = 1.31, 95%CI:1.18 - 1.45, *P* < 0.001). **(Table 2**)

**Figure 1.**
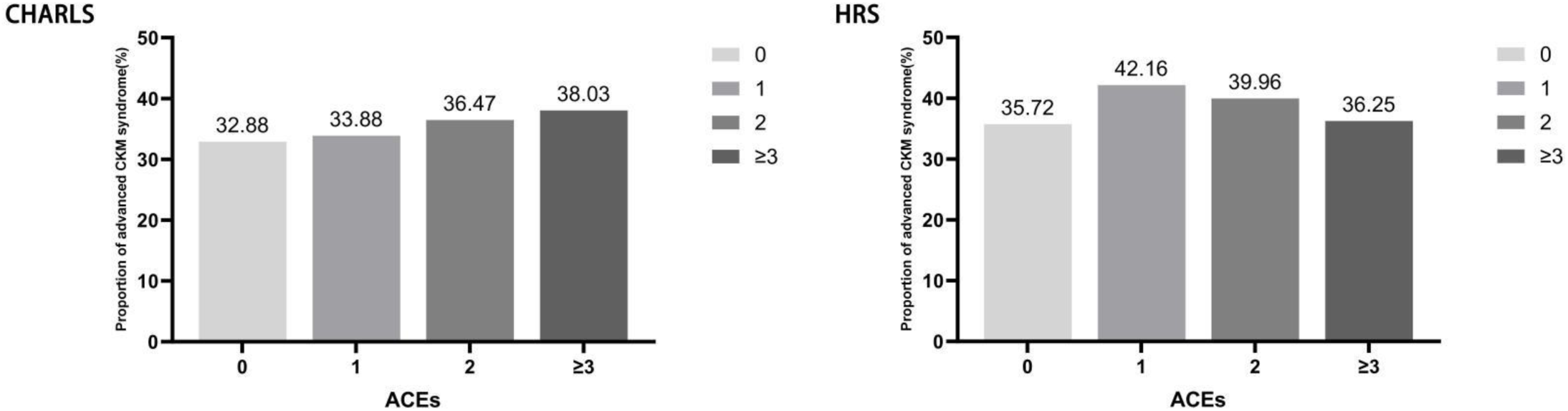
Proportion of CKM syndrome (stages 3 - 4) in participants with ACEs. ACEs: adverse childhood experiences; CKM: cardiovascular-kidney-metabolic syndrome CHARLS: China Health and Retirement Longitudinal Study; HRS: Health and Retirement Study.

**Table 2.** Association between ACEs and CKM syndrome.

| Number<br>of ACEs | CHARLS |  | HRS |  | Pooled |  | I<sup>2</sup> | Cochran's Q |
| --- | --- | --- | --- | --- | --- | --- | --- | --- |
|  | OR (95 % CI) | *P* value | OR (95 % CI) | *P* value | OR (95 % CI) | *P* value |  |  |
| Model 1 |  |  |  |  |  |  |  |  |
| 0 | Reference |  | Reference |  | Reference |  |  |  |
| 1 | 1.05(0.93,1.17) | 0.435 | 1.31(1.15,1.50) | <0.001 | 1.17(0.94,1.45) | 0.156 | 83.6% | 6.10 |
| 2 | 1.17(1.04,1.32) | 0.009 | 1.20(1.03,1.39) | 0.020 | 1.18(1.08,1.30) | <0.001 | 0.0% | 0.07 |
| ≥3 | 1.25(1.11,1.42) | <0.001 | 1.02(0.88,1.18) | 0.758 | 1.13(0.93,1.38) | 0.217 | 76.9% | 4.33 |
| Model 2 |  |  |  |  |  |  |  |  |
| 0 | Reference |  | Reference |  | Reference |  |  |  |
| 1 | 1.03(0.91,1.15) | 0.666 | 1.30(1.12,1.51) | 0.001 | 1.15(0.92,1.45) | 0.225 | 82.7% | 5.78 |
| 2 | 1.17(1.03,1.33) | 0.013 | 1.46(1.23,1.74) | <0.001 | 1.30(1.04,1.61) | 0.019 | 75.4% | 4.06 |
| ≥3 | 1.29(1.14,1.47) | <0.001 | 1.42(1.20,1.68) | <0.001 | 1.34(1.21,1.48) | <0.001 | 0.0% | 0.8 |
| Model 3 |  |  |  |  |  |  |  |  |
| 0 | Reference |  | Reference |  | Reference |  |  |  |
| 1 | 1.04(0.93,1.17) | 0.488 | 1.23(1.06,1.44) | 0.007 | 1.12(0.95,1.32) | 0.168 | 66.1% | 2.95 |
| 2 | 1.19(1.05,1.35) | 0.007 | 1.35(1.13,1.61) | 0.001 | 1.25(1.11,1.41) | <0.001 | 22.9% | 1.30 |
| ≥3 | 1.34(1.18,1.52) | <0.001 | 1.26(1.06,1.49) | 0.008 | 1.31(1.18,1.45) | <0.001 | 0.0% | 0.32 |

Model 1: unadjusted;
Model 2: adjusted for age and sex;
Model 3: adjusted for age, sex, marital status, education, smoking, drinking, race, and residential area;
CHARLS: China Health and Retirement Longitudinal Study; HRS: Health and Retirement Study; Statistical analyses were conducted using logistic regression.
Pooled effects were determined through random-effects meta-analysis.

### 3.3 Mediation analysis

As shown in **Figure 2**, the mediating effects of social support on the associations between ACEs and CKM syndrome were 2.1% and 11.8% in the CHARLS and HRS cohorts, respectively.

**Figure 2.**
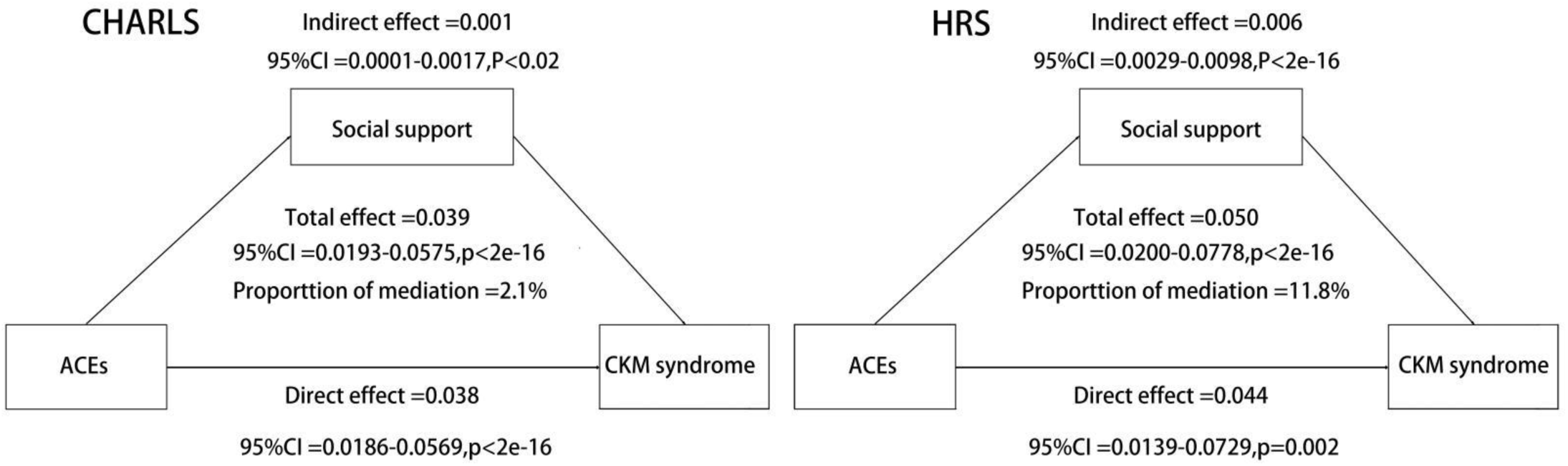
Mediating effects of social support in ACEs and CKM. CHARLS: China Health and Retirement Longitudinal Study; HRS: Health and Retirement Study; ACEs: adverse childhood experiences; CKM: Cardiovascular-Kidney-Metabolic Syndrome; Adjusted for age, sex, marital status, education, smoking, drinking, race, social support, and residential area; A total of 548 and 2749 participants were excluded due to missing data on social support in the CHARLS and the HRS cohort, respectively.

### 3.4 Subgroup analyses and sensitivity analyses

The results of subgroup analysis are presented in **Figure 3**, with no significant interaction effect being observed except for age and ACEs on CKM syndrome in the CHARLS (*P*_interaction_ = 0.022). In the sensitivity analysis, the associations of ACEs with physical abuse, being bullied, parental disability, domestic violence, death of a parent, separation from and parent were also consistent with the main analysis. **(Supplementary Table S3 and Table S4)** When ACEs was treated as a continuous variable (a linear regression model was used), the results were also consistent. **(Supplementary Table S5)**

**Figure 3.**
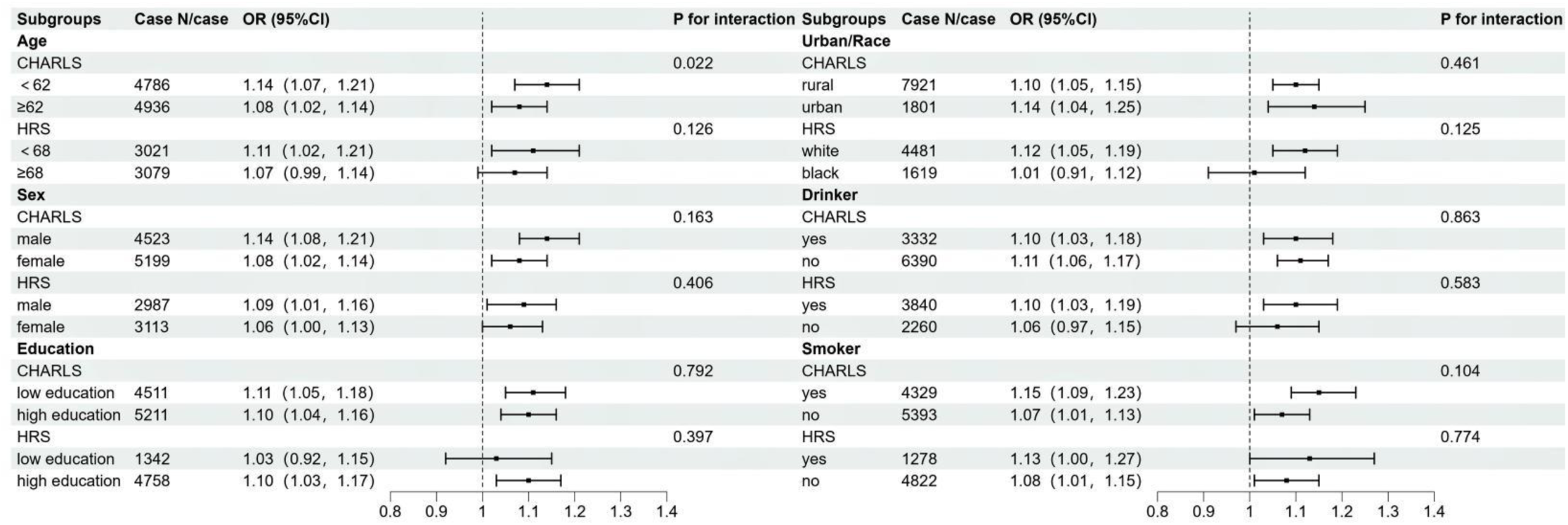
Subgroup analyses. CHARLS: China Health and Retirement Longitudinal Study; HRS: Health and Retirement Study. Statistical analyses were conducted using logistic regression, adjusting for age, sex, marital status, education, smoking, drinking, race, and residential area. *P* for interaction was assessed by adding multiplicative interaction terms between these factors and ACEs in the logistic regression models, with a significance level of 0.05. Values were presented as OR (95% CI), where OR represented the estimated odds ratio values and CI indicated the 95% confidence interval around the estimate. The small squares represent the odds ratio values, and the error bars represent the 95% confidence intervals for the odds ratio in each subgroup.

## 4 Discussion

In this pooled analysis, we examined the association between ACEs and CKM syndrome based on two national cohorts from China and the US. We found a positive association between ACEs exposure and CKM syndrome in the general population. Furthermore, our results suggest that social support plays a partial mediating role in this relationship.

Epidemiological studies have been supporting the important role of ACEs on health, including chronic kidney disease, premature mortality, CVD, and early onset of chronic conditions.^24-27^ Moreover, previous studies have shown the associations between ACEs and risk factors for those diseases, such as smoking, alcohol consumption,^28,29^ and stress.^30^ The AHA has also published scientific statements acknowledging ACEs as a social determinant of CVD risk and cardiometabolic health outcomes.^31^ Although the association between ACEs and some CKM components is well established, its role in CKM syndrome remains unclear. This study extends prior research by examining ACEs and CKM syndrome and indicates that elevated ACEs are associated with a higher risk of CKM syndrome.

Previous evidence suggests that early life social support may facilitate the development of effective coping and emotion regulation strategies to mitigate the impact of childhood and adulthood stress, as well as reduce CVD risk factors associated with ACEs.^32^ Additionally, the study result emphasizes that social support and belonging as potential intervention targets for reducing mental health risk among students with ACEs or other traumatic experiences.^33^ To further explore the potential role underlying the association between ACEs and CKM syndrome, we conducted a mediation analysis and reported that the proportion of the mediating effect of social support in the impact of ACEs on the incidence of CKM syndrome was 2.1% and 11.8%, respectively. The differences arising from Eastern and Western cultures, along with the resulting variations in social support forms, may explain the disparities in the mediating effects observed between the two cohorts.

The pathophysiology of CKM syndrome is characterized by a complex interplay of hemodynamic and neurohormonal mechanisms, including sympathetic overactivity, the renin-angiotensin-aldosterone system, various chemical mediators (nitric oxide, prostaglandins, endothelins, etc.), and oxidative stress.^34,35^ Epidemiological studies have consistently demonstrated that cardiovascular, renal, and metabolic diseases often overlap and coexist in the same patients. The mechanism linking ACEs to CKM syndrome involves a complex, multisystem network. As the initiating stressor, ACEs permanently disrupt stress response systems, particularly the hypothalamic-pituitary-adrenal axis and the sympathetic nervous system, resulting in dysregulated cortisol secretion and aberrant inflammatory responses.^36,37^ Chronic inflammation represents a central pathway in the onset and progression of CKM. In stage 1 CKM, excess and/or dysfunctional adiposity is the predominant feature, typically accompanied by adipocyte-driven low-grade inflammation.^38^ In stage 2, sustained metabolic risk factors amplify inflammatory signaling by recruiting additional immune cells, thereby perpetuating chronic inflammation. At this stage, insulin resistance and endothelial dysfunction emerge as key pathological processes, closely linked to immune cell-mediated oxidative stress.^39^ The progression of chronic kidney disease further intensifies inflammation, as early renal injury triggers lymphocytes, including B cells, to release proinflammatory mediators that promote macrophage infiltration and accelerate renal fibrosis through damage-associated molecular patterns.^40^ In advanced CKM, impaired cardiac and renal function leads to the accumulation of inflammatory mediators such as interleukin-6 and tumor necrosis factor-α, amplifying systemic inflammation.^41,42^. Concurrently, psychological trauma from ACEs fosters depression, anxiety, and maladaptive behaviors (e.g., poor diet, alcohol misuse), which interact with biological alterations in a bidirectional manner.^43,44^ Together, these psychosocial and biological pathways drive obesity, hypertension, diabetes, and dyslipidemia, which synergistically impair renal function and ultimately culminate in multi-organ metabolic failure, completing the pathological cascade from early psychological trauma to advanced CKM syndrome.

One major strength is that we for the first time investigated the association between ACEs and CKM, and the results are mutually verified in two nationally representative cohorts. Additionally, we explored the potential role of social support in the association. However, there are several limitations of our study as well. First, due to the research designs, the items of the ACEs questionnaires of the two cohorts were different, which resulted in the large heterogeneity of results and prevented us from exploring the effects of different categories of ACEs (such as conventional ACEs, expanded ACEs, and novel ACEs) on CKM. Second, the collection of ACE indicators relied on retrospective data, potentially introducing recall bias, particularly to subjective events like neglect. Third, out of sensitivity, privacy concerns, or psychological distress of the ACEs items, participants may decline to respond or intentionally provide incorrect answers to certain questions. Finally, we did not account for the frequency, severity, and duration of ACEs, factors known to correlate with adverse health consequences.

## 5 Conclusion

In summary, we for the first time reported that ACEs were significantly associated with CKM syndrome in the general population. Furthermore, our results suggest that social support plays a partial mediating role in this relationship. Our research provided more insights into the role of ACEs on adverse health outcomes and underscored the important role of social supports in the association; however, future research is needed to identify which forms of social support are effective across different cultural contexts.

## Data Availability

no

https://charls.pku.edu.cn/en/

https://hrs.isr.umich.edu/

## Sources of Funding

The authors declare that no funds, grants, or other support were received during the preparation of this manuscript.

## Disclosures

None.

## Supporting information

Tables S1-S5 Figures S1-S2

## Acknowledgements

We sincerely thank all staffs and respondents related to the HRS and the CHARLS for their effort and contribution.

## CRediT authorship contribution statement

**Xiu Qin:** Writing - original draft, Visualization, Software, Formal analysis. **Haofeng Zhang:** review & editing. **Fudong He:**review & editing. **Lei Sun:**review & editing. **Hua Xiao:**review & editing. **Yufan Nan:**review & editing. **Haiyan Chen:**review & editing. **Haidong Zhu:**review & editing. **Yanbin Dong:**review & editing. **Guang Hao:** Writing - review & editing, Supervision, Conceptualization.

---

**Supplementary Table S1.** Details of ACEs of two cohorts.

| Supplementary Table S1. Details of ACEs of two cohorts. |  |  |  |  |
| --- | --- | --- | --- | --- |
| No. | Abbreviation | Items | Question | Answer is defined as exposure |
| HRS |  |  |  |  |
| 1 | PE | Parental education | What is the highest grade of school your father/mother completed? | No formal education, Grades |
| 2 | PAP | Physical abuse | Before you were 18 years old, were you ever physically abused by your parents? | Yes |
| 3 | PAD | Parental alcohol and drug use | Before you were 18 years old, did either of your parents drink or use drugs so often that it caused problems in the family? | Yes |
| 4 | SFP | Separation from parents | Were you ever separated from your mother/father for 6 months or longer? | Yes |
| 5 | DP | Death of a parent | Did one or both of your biological or adoptive parents die? | Yes |
| 6 | RCO | Residing in a children's home or orphanage | Did you ever live in a children's home or orphanage? | Yes |
| 7 | CP | Childhood poverty | Would you say your family during that time was pretty well off financially, about average, or poor? | Poor |
| 8 | PSD | Parental separation or divorce | Did your biological or adoptive parents separate or divorce? | Yes |
| CHARLS |  |  |  |  |
| 1 | PAP | Physical abuse | When you were growing up, did your female/male guardian ever hit you? | Often/Sometimes |
| 2 | EN | Emotional neglect | How much love and affection did your female guardian give you while you were growing up? | Rarely/Never |
| 3 | SAH | Substance abuse within the household | During the years you were growing up, which one of the followings did your female/male guardian ever have? | Alcoholism/Drug |
| 4 | MIH | Mental illness in the household | Did your female/male guardian have abnormality of mind when you were young? | Yes |
| 5 | DV | Domestic violence | Have your father ever beat up your mother?/Have your mother ever beat up your father? | Often/Sometimes |
| 6 | IHM | Having an incarcerated household member | During the years you were growing up, which one of the followings did your female/male guardian ever have? | Arrested or sent to prison |
| 7 | PSD | Parental separation or divorce | Were your biological parents divorced? | Yes |
| 8 | LUN | Living in an unsafe neighborhood | Was it safe being out alone at night in the neighborhood where you lived as a child? | Not very safe/Not safe at all/ Not very often Being bullied |
| 9 | BB | Being bullied | When you were a child, how often were you picked on or bullied by kids in your neighborhood/school? Is it often, sometimes, rarely or never? | Often/Sometimes |
| 10 | DP | Death of a parent | At least one of the participant's parents had passed away before they were 17. | $\geq 1$ |
| 11 | DS | Death of a sibling | One of the participant's siblings had passed away before they were 17 years. | $\geq 1$ |
| 12 | PD | Parental disability | Did your female/male guardian have a long time be sick on bed when you were young? Yes Did your female/male guardian have a serious deformity when you were young? | Yes |

\*Items calculated according to the birthdays of participants, their parents, and siblings.
CHARLS: China Health and Retirement Longitudinal Study; HRS: Health and Retirement Study.

**Supplementary Table S2.** Details of the social support in the two cohorts.

|  |  |  |
| --- | --- | --- |
| HRS |  |  |
| 1 | How much do they really understand the way you feel about things? | 1 (a lot), 2 (some), 3 (a little), 4 (not at all) |
| 2 | How much can you rely on them if you have a serious problem? | 1 (a lot), 2 (some), 3 (a little), 4 (not at all) |
| 3 | How much can you open up to them if you need to talk about your worries? | 1 (a lot), 2 (some), 3 (a little), 4 (not at all) |
| CHARLS |  |  |
| 1 | When you were a young adult, was there anyone who provided you with financial support for your work? | Yes<br>No |
| 2 | When you were a young adult, was there anyone who provided you with positive nonfinancial support for work? | Yes<br>No |
| 3 | When you were a young adult, was there anyone who provided you with positive support or mentoring for your interpersonal relationship? | Yes<br>No |

**Supplementary Table S3.** Association between ACEs and CKM syndrome in the CHARLS cohort.

| Types of ACEs | OR (95 % CI) | *P* value |
| --- | --- | --- |
| Being bullied | 1.18(1.04,1.34) | 0.010 |
| Physical abuse | 1.25(1.13,1.38) | <0.001 |
| Death of a parent | 1.05(0.92,1.19) | 0.484 |
| Death of a sibling | 1.09(0.95,1.25) | 0.208 |
| Parental disability | 1.32(1.19,1.47) | <0.001 |
| Emotional neglect | 1.00(0.91,1.10) | 0.926 |
| Domestic violence | 1.12(1.00,1.25) | 0.050 |
| Living in an unsafe neighborhood | 1.08(0.92,1.27) | 0.350 |
| Parental separation or divorce | 1.28(0.80,2.04) | 0.302 |
| Mental illness in the household | 1.15(0.91,1.44) | 0.237 |
| Substance abuse within the household | 0.97(0.81,1.16) | 0.748 |
| Having an incarcerated household member | 1.03(0.43,2.42) | 0.953 |

adjusted for age, sex, marital status, education, smoking, drinking, race, and residential area;
ACEs: adverse childhood experiences;
CHARLS: China Health and Retirement Longitudinal Study; HRS: Health and Retirement Study;
Statistical analyses were conducted using logistic regression.

**Supplementary Table S4.** Association between ACEs and CKM syndrome in the HRS cohort.

| Types of ACEs | OR (95 % CI) | *P* value |
| --- | --- | --- |
| Physical abuse | 1.47(1.20,1.80) | <0.001 |
| Childhood poverty | 1.10(0.96,1.26) | 0.162 |
| Death of a parent | 1.27(1.10,1.46) | 0.001 |
| Separation from parents | 1.14(1.00,1.31) | 0.044 |
| Parental alcohol and drug use | 1.09(0.94,1.26) | 0.258 |
| Parental separation or divorce | 1.07(0.92,1.26) | 0.372 |
| Residing in a children’s home or orphanage | 1.00(0.66,1.51) | 0.994 |

adjusted for age, sex, marital status, education, smoking, drinking, race, and residential area;
ACEs: adverse childhood experiences;
CHARLS: China Health and Retirement Longitudinal Study; HRS: Health and Retirement Study;
Statistical analyses were conducted using logistic regression.

**Supplementary Table S5.** Association between ACEs and CKM syndrome.

| Number of ACEs | CHARLS |  | HRS |  |
| --- | --- | --- | --- | --- |
|  | Beta (95 % CI) | *P* value | Beta (95 % CI) | *P* value |
| Model 1 |  |  |  |  |
| 0 | Reference |  | Reference |  |
| 1 | -0.02(-0.08,0.04) | 0.582 | 0.14(0.06,0.21) | <0.001 |
| 2 | 0.05(-0.01,0.12) | 0.118 | 0.10(0.01,0.18) | 0.025 |
| ≥3 | 0.05(-0.01,0.12) | 0.123 | 0.03(-0.05,0.11) | 0.524 |
| Model 2 |  |  |  |  |
| 0 | Reference |  | Reference |  |
| 1 | -0.02(-0.08,0.04) | 0.516 | 0.10(0.04,0.17) | 0.002 |
| 2 | 0.06(-0.01,0.12) | 0.079 | 0.17(0.09,0.24) | <0.001 |
| ≥3 | 0.07(0.01,0.14) | 0.034 | 0.17(0.10,0.25) | <0.001 |
| Model 3 |  |  |  |  |
| 0 | Reference |  | Reference |  |
| 1 | -0.01(-0.07,0.05) | 0.774 | 0.08(0.02,0.15) | 0.015 |
| 2 | 0.07(0.01,0.13) | 0.033 | 0.13(0.05,0.21) | 0.001 |
| ≥3 | 0.10(0.03,0.16) | 0.004 | 0.12(0.05,0.19) | 0.001 |

Model 1: unadjusted;
Model 2: adjusted for age and sex;
Model 3: adjusted for age, sex, marital status, education, smoking, drinking, race, and residential area;
ACEs: adverse childhood experiences;
CHARLS: China Health and Retirement Longitudinal Study; HRS: Health and Retirement Study;
Statistical analyses were conducted using linear regression.

**Supplementary Figure 1.**
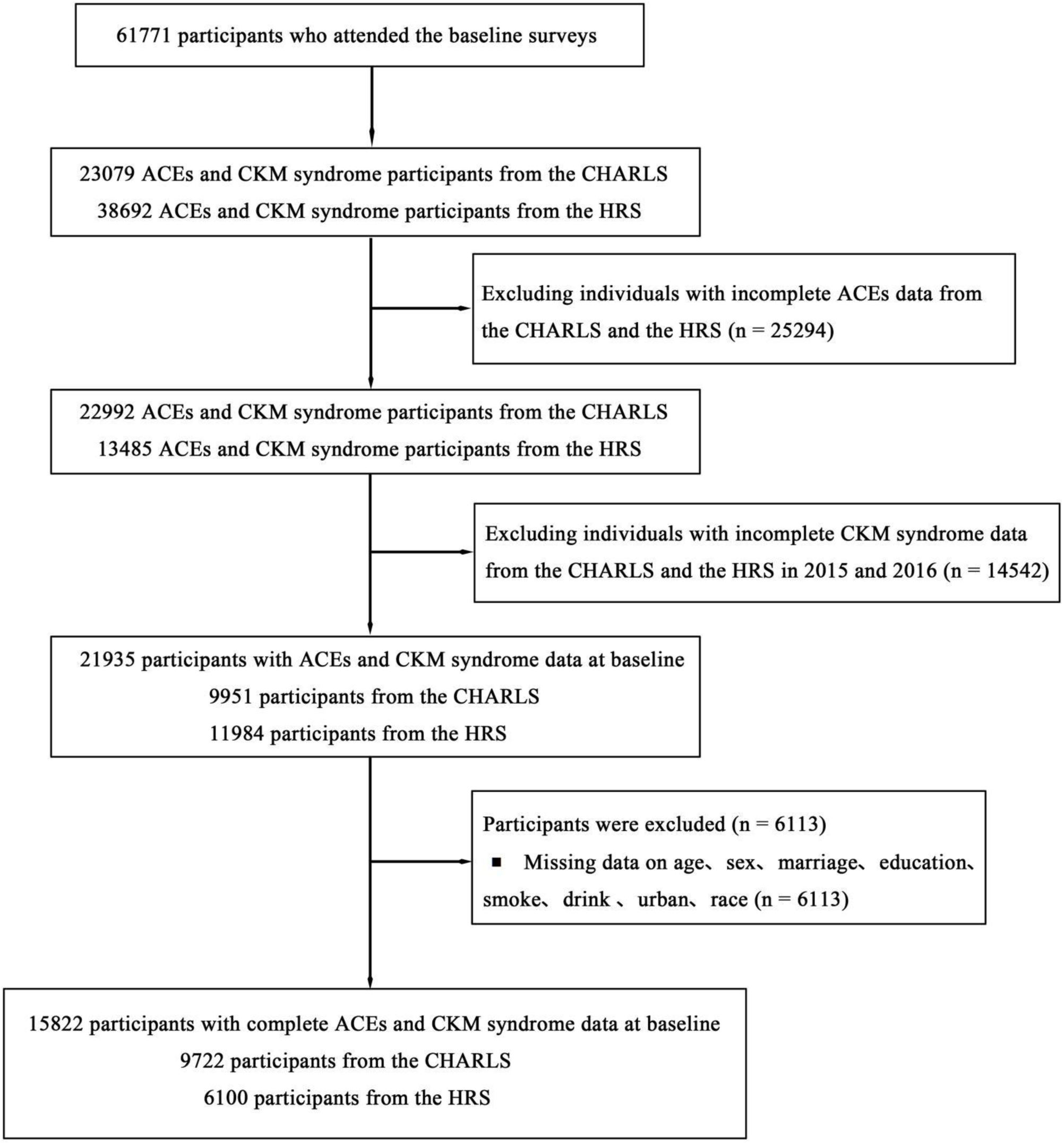
The flow chart of study participants.

**Supplementary Figure 2.**
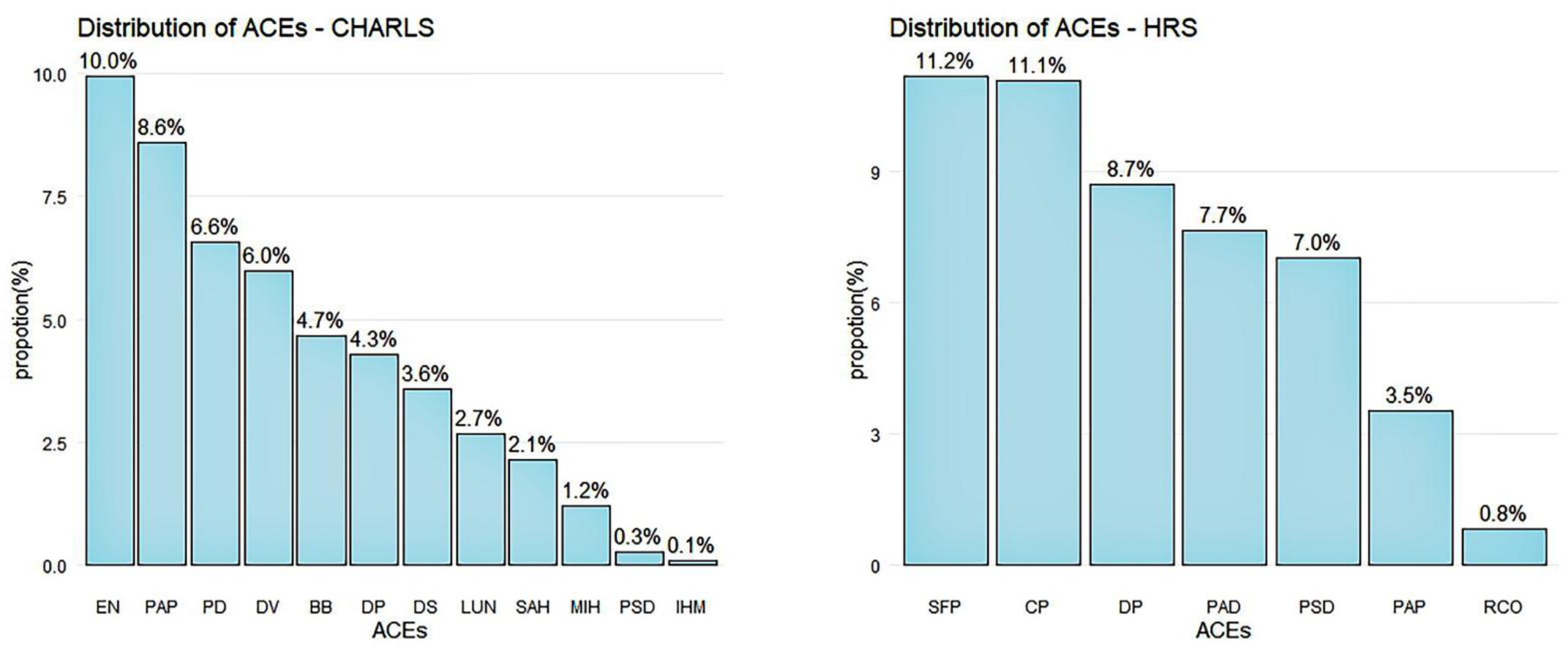
The distribution of ACEs among two cohorts. HRS: Health and Retirement Study; CP: Childhood poverty; SFP: Separation from parent; DP: Death of a parent; PAD: Parental alcohol and drug use; PSD: Parental separation or divorce; PAP: Physical abuse; RCO: Residing in children’s home or orphanage. CHARLS: China Health and Retirement Longitudinal Study; EN: Emotional neglect; PAG: Physical abuse by guardian; PD: Parental disability; DV: Domestic violence; BB: Being bullied; DP: Death of a parent; DS: Death of a sibling; LUN: Living in an un safe neighborhood; SAH: Substance abuse within the household; MIH: Mental illness in the household; PSD: Parental separation or divorce; IHM: Having an incarcerated household membe

## Notes

### Competing Interest Statement

The authors have declared no competing interest.

### Clinical Trial

no

### Author Declarations

This study was performed in line with the principles of the Declaration of Helsinki and all participants signed informed consents before participation. Approval was granted by the Ethical Review Committee at Peking University (IRB00001052?11015)

